# The Effects of Prenatal Deaths on National Life Expectancy: Case Study U.S.A

**DOI:** 10.1101/2023.03.06.23286890

**Authors:** Madeleine R. Hollman, Joshua M. Pearce

## Abstract

It is a positive indicator that human life expectancies calculated from birth have been increasing. The current standards for counting life-years, however, assume social desirability and exclude all prenatal deaths. These assumptions mask low life-year deaths and obscure results of medical and environmental interventions, thus falsely indicating higher life expectancies. This case study investigates 1930 to 2016 using CDC and World Bank data for the U.S. It is evident, published U.S. life expectancies are greatly exaggerated and what would have been short-lived Americans are disproportionately labeled as socially-undesirable and ignored when counting life years, thus presenting an overly-optimistic view of U.S. health. A comprehensive global investigation is needed, and a refinement of life expectancy calculations should be introduced, which does not bias results by only counting life expectancy from the time of live birth.

## Introduction and Background

Both globally and at the national scale, human life expectancies have been increasing [1,2]. This positive social outcome is generally attributed to improvements in hygiene, education (particularly that of women), and improvements in medical science and technology [2]. The current international standards for counting life-years from live birth to over 100 years assumes a subjective social desirability (e.g. only the life times of humans that society has determined to be socially desirable are included in the calculations) [3,4]. This assumption is carried through all subsequent analysis such as the disability adjusted life year (DALY) that are used to drive health policy [5,6]. This assumption, however, may mask low life-year deaths caused by socioeconomic or other considerations, and thus may provide incorrect life expectancies in a given population. In addition, ignoring prenatal deaths may mask the impacts of medically-relevant environmental factors, priority areas for intervention, and health policies that would otherwise influence the longevity for a given population. In order to obtain a complete, in-depth understanding of national health gains, this preliminary investigation provides a methodology to determine the impact on national life expectancy statistics when both socially-undesirable individuals and prenatal deaths are included. A case study is performed using this methodology for the United States (U.S.). The results are discussed in the context of optimizing national and international health policy.

## Methods

An 86 year period (1930 to 2016) is investigated using the U.S. Center for Disease Control (CDC) data and extended by the World Bank comparing national life expectancy calculations taking into account social desirability from the CDC and the Guttmacher Institute [4,7-10]. Birth counts are found using CDC data and are extended with data from the U.S. Department of Health and Human Services [7,11]. To include pregnancy terminations into life expectancies, the following equation is used for the corrected life expectancy (L_CSD_):

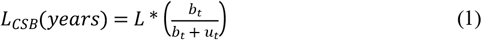

Where L is the standard life expectancy, b_t_ is the number of socially-desirable births/year in a given year t, and u_t_ is the number of pregnancy terminations in year t.

To account for miscarriages per year at year t (m_t_) equation (1) is adjusted with data from the National Center for Health Statistics (NCHS) by the CDC [12].

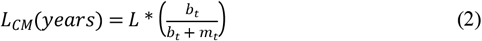

To combine both pregnancy terminations and miscarriages into the corrected life expectancy, the corrected life expectancy, L_c_, becomes:

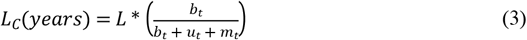

To assess life expectancy if terminated pregnancies had not been terminated, the data is divided by race/ethnicity and poverty level. The rate of pregnancy termination is higher in minority races and ethnicities as well as in people below the federal poverty line [4,13]. To calculate life expectancy if terminated pregnancies had resulted in individuals who had lived to their respective projected life expectancy, L_c_, becomes:

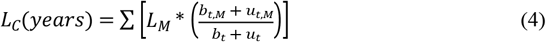

where b_t,M_ is the number of socially-desirable births per year in a given year t for a racial group, M, and u_t,M_ is the number of pregnancy terminations in year t per group [9,11,12,14]. The racial groups included in this calculation are Hispanic, African American, Caucasian and other, each with a life expectancy L_m_. Equation (4) is also used to account for the population above and below the federal poverty line [13,15,16]. Due to the statistics of pregnancy terminations from the CDC not being comprehensive of the U.S. population, to proportionally estimate the number of lives terminated per racial group [17]:

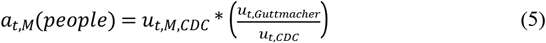

The variables include u_t,m,CDC_ being the number of pregnancies terminated per racial/ethnic group, according to the incomplete CDC data, u_t,CDC_ is the total pregnancies terminated according to the CDC, and u_t,Guttmacher_ is the total pregnancies terminated according to the more thorough Guttmacher Institute data, all in year t.

The most comprehensive life expectancy data correlating income and life expectancy from Chetty et al. divides the population by percentile in 2001-2014 [15]. In this calculation, the bottom 14% are considered below the federal poverty line. The calculation to find life expectancy above and below the federal poverty line is:

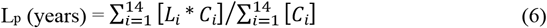

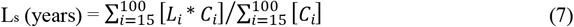

where L_p_ is the life expectancy below the federal poverty line and L_s_ is the life expectancy above the federal poverty line. C_i_ is the count of people in each percentile and L_i_ is the life expectancy of the people in each percentile. The overall life expectancy of this study is found to be higher than that of the CDC, so for comparison, the values are adjusted for the corrected life expectancy, L_p,CDC_ :

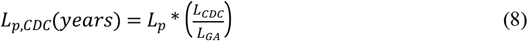

where L_p,CDC_ is the life expectancy adjusted to the CDC data, L_CDC_ is the total life expectancy from the CDC and World Bank, and L_GA_ is the total life expectancy using Gompertz Approximation to mortality rates in 2001 NCHS data [15].

## Results and Discussion

Traditionally-calculated life expectancy is compared to the life expectancy of the U.S. including miscarriages and those pregnancy terminations in Figure 1. It is clear from Figure 1 that when miscarriages are included (blue diamonds), the rise in U.S. life expectancies are significantly muted (about 2 years of increased life time in 35 years). This is in stark contrast to the conventional calculation (green filled circles), which is more than double that rate. The causes of increasing miscarriage rates are heavily disputed and there are many possible causes that would be expected to increase the rate. The trend is seen especially in early term miscarriages, which may be due to increased reporting, likely due to increased awareness [18]. Contraceptive use has also increased (i.e. in 2011-2015 15.0% of U.S. women used an IUD and 79.3% used birth control pills [19]) and causes early term miscarriages [19]. If pregnancy occurs with an IUD in place, miscarriage occurs in 50% of cases where the IUD is left in place and 25% of cases where the IUD is subsequently removed [20]. Furthermore, IUDs are associated with actinomycosis and tubular pregnancy, which often lead to miscarriage [20,21]. Even after the IUD has been removed, women who have had them for more than three years were found to be twice as likely to have tubal pregnancy [21]. The use of oral contraceptives for more than two years has also been associated with higher risk of miscarriage [22]. The termination of pregnancy by vacuum aspiration increases the risk of first-trimester miscarriages as well [23]. There are also potential external factors that could contribute to miscarriages, such as chemical pollution, pesticide exposure, nutritional deficiencies, infections, hormonal imbalances, menstrual disorders, psychological trauma, and stress life events [24,25].

**Figure 1.**
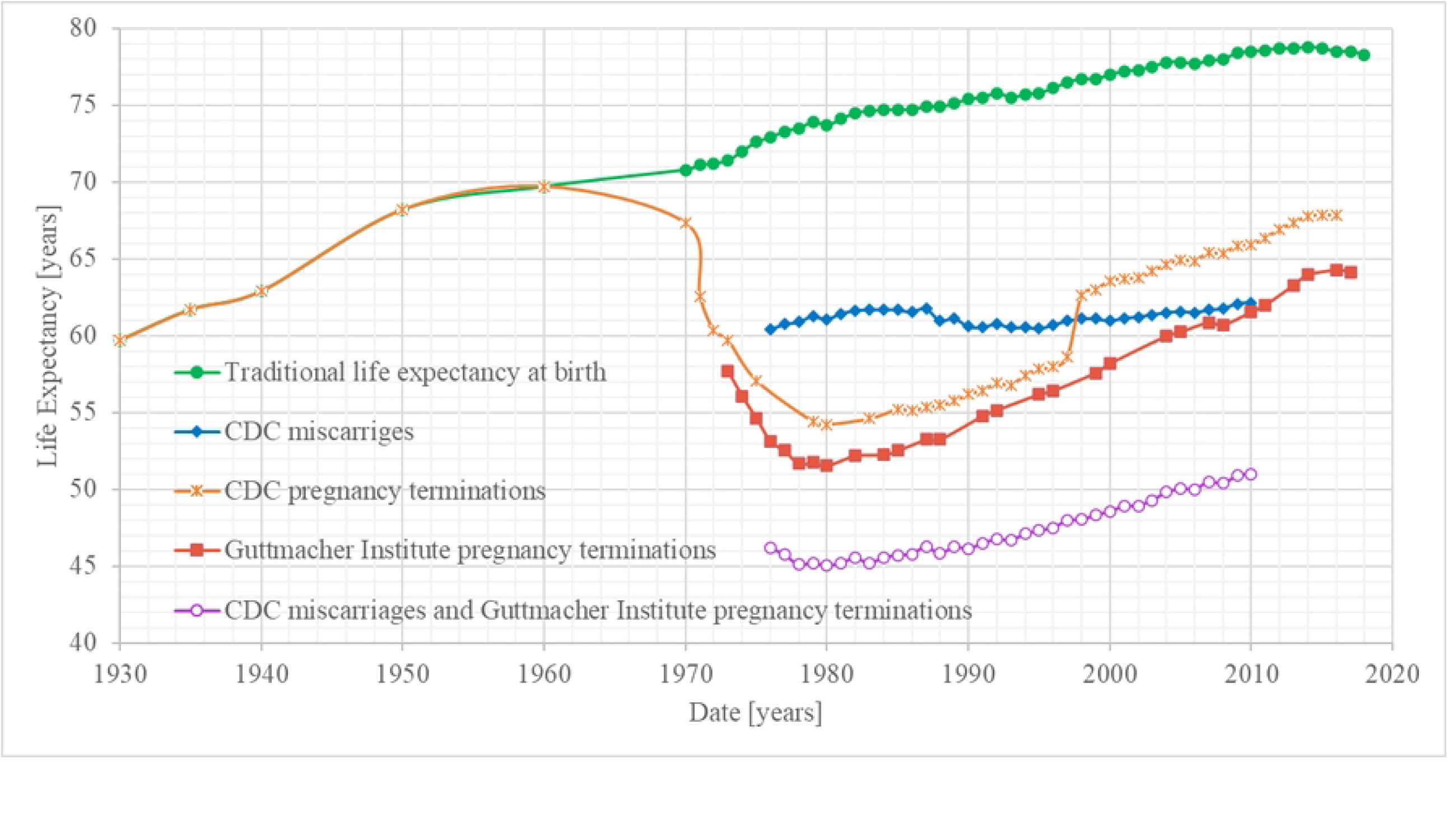
Life expectancies from 1930 to 2016 as a function of years with and without social undesirables and miscarriages included.

Traditional life expectancy in the United States has increased from 59.7 to 78.3 years from 1930 to 2018 (green filled circles in Figure 1). This is an 18.6-year increase in life expectancy. Including the CDC’s count of pregnancy terminations, the life expectancy does not follow a monotonic traditional increasing trend. The primary change in the corrected life expectancy shown in Figure 1 (orange x) is due to the legalization of prenatal termination in the U.S. in 1973. It can also be seen that the CDC data makes a jump between 1997 and 1998 because four states (Alaska, California, New Hampshire, and Oklahoma) stopped reporting this data to the CDC, which is voluntary [26]. Furthermore, as of 2020, five states (Alabama, Florida, Hawaii, Nevada, and Tennessee) only report surgical (not medical) pregnancy terminations, two states (California, Maryland) have no reporting requirements, and in three states (New Hampshire, New Jersey, and DC), it is voluntary for physicians to report [17]. The Guttmacher Institute has been collecting data since 1973 and is the primary source used in U.S. policy due to its greater accuracy by overcoming these complications of data collection from government sources [25]. Life expectancy including the count of pregnancy terminations from the Guttmacher institute (red filled squares) is below the CDC outcome (orange x) with a more consistent trend. In 1996, the Guttmacher Institute reported 138,575 more terminations, resulting in 1.5 less years of life expectancy than the CDC had reported, and in 1999, they reported 452,991 more terminations, causing 5.4 less years in the life expectancy calculation.

The greatest difference in traditional life expectancy to the life expectancy including pregnancy terminations is 20.0 years in 1983. The smallest difference is in 2016, with a 10.7-year gap. Including miscarriages reduces the life expectancy by 12.5-16.4 years over the entire range where data was available. Due to the increase in reported miscarriage rates, the greatest difference in life expectancy is with the most recent data in 2010 [12]. The lowest life expectancy line (purple open circles) includes both miscarriages and pregnancy terminations taken into account. In 1976, this reduces life expectancy by 26.7 years from the traditional 72.9 years to 46.2 years. With the most recent available data, in 2010 including pregnancy terminations and miscarriages reduces life expectancy by 27.5 years from 78.5 to 51.0 years. The greatest difference in life expectancy is 29.4 years.

In addition to rough national statistics shown in Figure 1, Table 1 probed the impact of socioeconomic status and race on life expectancy when accounting for pregnancy termination. If these individuals lived a full life, life expectancy was found to be lower. When separating people by ethnicity and race, however, there is not a significant difference in life expectancy by including this correction. When separating the population above and below the poverty line, life expectancy, however, would be lower if the terminated pregnancies had come to term and had lived to full life expectancy. This phenomenon is because people in poverty have a lower life expectancy and higher rate of prenatal termination [13,28].

**Table 1.**
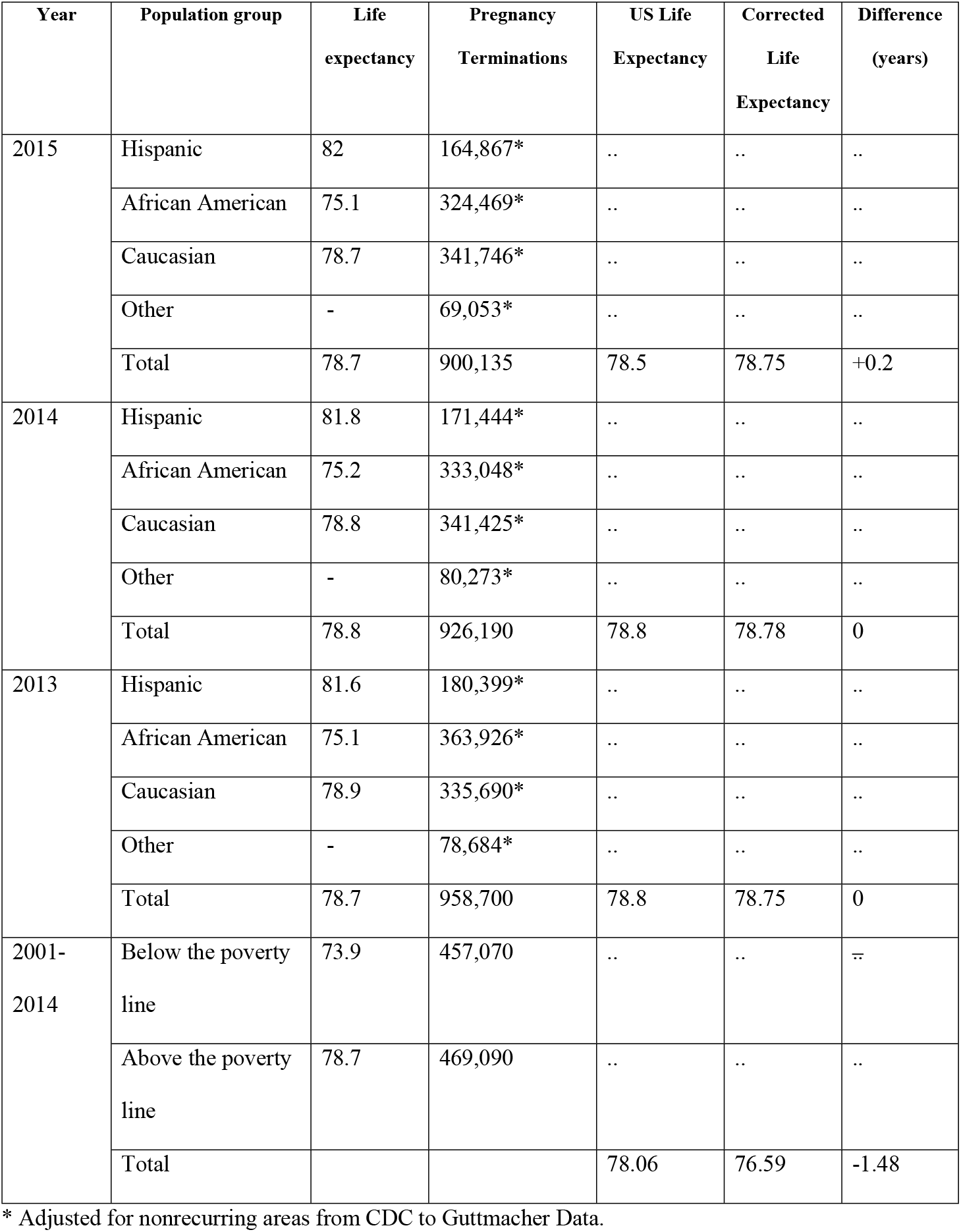
Socioeconomic status correction on life expectancy in the U.S.

It is evident from the results of this study that there is a great need for more complete and comprehensive data collection to gain a more thorough understanding of life expectancy in the U.S. Information, when it is available, is misleading and far from comprehensive. This approach can also be used in other countries to gain a better understanding of real life expectancies. Different countries have vastly different and changing laws, which will make future work with cross-country comparisons challenging. Despite the known challenges, this is important because varying laws and social practices in different countries would be expected to influence the traditional life expectancy estimation, making the current comparisons incorrect. For example, consider only the case of Down syndrome, which has a traditional life expectancy of about 60 years, which is still substantially lower than the traditional 78.5 years estimated for the whole U.S. population. Future work, is needed to make the correction demonstrated in this article for life expectancy and compare the U.S., which has a 67% termination rate for pregnancies that have tested positive for Down syndrome with other countries with varying rates (e.g. 77% in France, 98% in Denmark, and 100% in Iceland) [29,30].

Only by recording correct life expectancies, including all deaths with complete data, can the global medical community hope to enact informed medical policies. A comprehensive data set will show more clearly what the causes of increasing miscarriages in the U.S. are and help reign in the current debate. Most importantly, improved medical policy based on comprehensive life expectancy calculations would be expected to help optimize medical care and nutrition, reduce future miscarriages and improve metal health, reduce insurance costs, and help develop appropriate environmental regulations to reduce toxic chemicals and pollutants that impact life expectancy.

## Conclusions

Life expectancy calculations, including pregnancy terminations, show up to 27.5 year gaps in perceived life expectancy in the U.S. and the reality. Published U.S. life expectancies are distorted due to short-lived citizens not being included in calculations because they are arbitrarily considered socially undesirable. Prenatal termination rates are decreasing since peaking in 1983, but miscarriage rates are increasing. The cause for increasing miscarriages is not well determined, but an increase in reporting, contraceptive use, and previous fetal terminations are likely. Including miscarriages and pregnancy terminations, the life expectancy in the U.S. is 51 years (27.5 years less in 2010 than to the CDC reported 78.5 years). If those pregnancies terminated had come to term and lived a full life, the life expectancy would be 1.5 years lower due to low income people having higher termination rates and lower life expectancies.

The results of this article make it clear that a redefinition of who is included in life expectancy calculations is needed in the U.S. as well as globally to remove the arbitrary bias of not including prenatal deaths in these calculations. To improve the health and well-being of the U.S. and global populations, optimal health policies, supported by more accurate, up-to-date, and publicly-accessible data about all pregnancies is essential for guiding national and international laws.

## Data Availability

All relevant data are within the manuscript and its Supporting Information files.

